# Bacterial etiologies, antimicrobial susceptibility profiles and associated factors among patients with otitis media referred to Nekemte Public Health Research and Referral Laboratory Center, Western Ethiopia: A prospective cross-sectional study

**DOI:** 10.1101/2024.04.02.24305211

**Authors:** Endalu Tesfaye Guteta, Fedassan Alemu Abdi, Seifu Gizaw Feyisa, Belay Merkeb Zewudie, Betrearon Sileshi Kinfu, Hunduma Feyisa Geleta, Tadesse Bekele Tafesse

## Abstract

**Background:** Otitis media is among the leading causes of illnesses responsible for causing hearing problems and adding significant costs to the public health system. Bacteria are the most common causative agents for otitis media. Currently, there is little information on the prevalence and antimicrobial susceptibility patterns of pathogenic bacterial isolates from patients with otitis media in Ethiopia.

**Methodology:** A laboratory – based cross-sectional study was conducted from June to September 2023 among 242 patients with otitis media referred to Nekemte Public Health Research and Referral Laboratory Center. Sociodemographic and clinical data were obtained by trained nurses and/or health officers in face-to-face interviews using structured questionnaires. Middle ear discharge samples were collected by trained clinical microbiology experts following all aseptic techniques. Conventional culture, different biochemical tests and antimicrobial susceptibility testing were performed for all the isolated bacteria. Reference strains were used as a positive and negative controls. The data were checked for completeness and consistency, entered into EpiData version 4.6.06 and analyzed by SPSS version 25. Logistic regression analysis was performed to determine the associated factors of otitis media. Adjusted odds ratio was used to determine strength of association. Statistical significance was obtained at p-value of below 0.05. The data were interpreted using graphs, tables, and results statements.

**Results:** A total of 242 middle ear discharge samples were collected and cultured from which 212 (87.6%) were culture positive. A total of 228 pathogenic bacterial isolates were recovered. The predominant bacterial isolates were *S. aureus* 92 (40.4%) followed by *P. aeruginosa* 33 (14.5%) and *E. coli* 24 (10.5%). One hundred fifty-one (66.2%) bacterial pathogens were multidrug resistant. Piperacillin-tazobactam and tobramycin are relatively common drugs to which most of the isolates were susceptible while ampicillin and tetracycline were the most resistant. Purulent discharge (p-value = 0.001), middle ear discharge ≥14 days (P-value = 0.000) and a history of active/passive smoking (P-value = 0.043) were significantly associated with otitis media.

**Conclusion:** The prevalence of bacterial pathogens, most of which were multidrug-resistant, was high among patients with otitis media. A significant association was observed with purulent ear discharge, chronic otitis media, and passive or active smoking. Choosing the proper antibiotic for the treatment of bacterial infection is crucial.

## Background

Otitis media (OM) is a common inflammatory illness that can affect people of all ages and results in temporary or permanent hearing loss due to fluid effusion or pathological alterations in the tympanic membrane of the middle ear (1). The spectrum of illnesses associated with OM ranges from acute to chronic and is clinically indicated by the presence of fluid accumulation in the middle ear that can cause temporary hearing loss (2).

Otitis media (OM) is a significant contributor to hearing loss in people of all ages but is more common among children under-five years of age. Globally, among 709 million patients with AOM, 51% were children under five years old. On the other hand, AOM is responsible for approximately 25% of antibiotic prescriptions among children (3–5). The peak prevalence of OM among children is mainly attributed to immature immune status, the anatomy of the Eustachian tube (shorter and horizontal), frequent exposure to URTIs and malnutrition (6,7).

OM is increasingly prevalent in Sub-Saharan Africa and other developing nations including Ethiopia. Due to limited microbiological laboratories, Sub-Saharan African nations rely solely on clinical data for therapy which could be responsible for more complicated OM (8). Bacteria are the most common agents for OM. The most predominant pathogenic bacteria isolated from AOM included *M. catarrhalis, S. pneumoniae* and *H. influenzae*. On the other hand, *P. aeruginosa*, *Klebsiella species*, *Proteus species*, *S. aureus* and *E. coli* are the predominant bacteria isolated from patients with CSOM (9).

The extensive application of antibiotics to treat OM has led to the growth of resistant bacteria, including strains that are resistant to many drugs (10). Several studies have reported considerable resistance rates of bacteria isolated from middle ear. To ensure the best possible care and fight antibiotic resistance, it is essential to continuously and periodically evaluate the microbiological profile and antimicrobial susceptibility (11,12).

In Ethiopia, few studies have investigated bacteriological profiles and antimicrobial susceptibility patterns of the middle ear discharge. The number of antibiotic-resistant bacteria is alarming and becoming a major public health problem in the management of patients with middle ear infection (13–15). There are no data on the prevalence and antimicrobial susceptibility patterns of bacterial pathogens from middle ear infections in referred health facilities. This on the other hand might indicate that clinicians base their treatment on empirical evidence. Knowing the local antibiogram is important for cost-effective and appropriate treatment of otitis media and helps prevent complications that may arise due to the lack of treatment or improper treatment. Thus, the aim of this study was to acquire data on bacterial pathogens responsible for otitis media and their antibacterial susceptibility patterns among patients with otitis media referred to the Nekemte Public Health Research and Referral Laboratory Center for culture tests.

## Methods

### Study area, study period and design

This study was conducted at Nekemte Public Health Research and Referral Laboratory Center among referred patients with otitis media from nearby health facilities from June to September 2023. A laboratory-based cross-sectional study was conducted in 242 patients with otitis media.

### Study population and sampling technique

Patients with middle ear discharge accompanied by laboratory-appropriate request form and those who were not receiving antibiotic were included in the study. Participants were selected through convenience sampling.

## Sample size

Sample size was calculated using a single population proportion formula by considering 80.4% prevalence of otitis media from study done at Bahir Dar Regional Health Research Laboratory Center (29), 95% confidence level, and 5% tolerable error (d = 0.05)

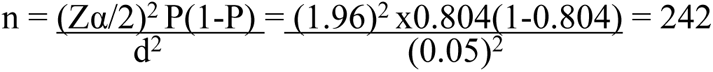

## Data collection, sample processing and laboratory analysis

Sociodemographic and clinical data were collected in face-to-face interviews from study participants by nurses/health officers using structured questionnaires. Middle ear discharge data were collected by trained clinical microbiology experts. The collected discharge samples were inoculated on blood agar, chocolate agar and MacConkey agar following standard bacteriological techniques. Bacterial pathogens from positive culture results were identified by their characteristic appearance on the respective media, gram-staining reaction and pattern of biochemical profiles using standard procedures.

The antibiotic susceptibility all the isolated bacterial pathogens was tested via Kirby-Bauer disc diffusion method. Inocula were prepared by transferring 3 - 5 colonies from pure culture into 5 ml of normal saline and mixing thoroughly to make a homogenous suspension equivalent to the 0.5 McFarland standard. Using a sterile cotton swab, the bacterial suspension was distributed evenly over the entire surface of MHA (HIMEDIA, India) and left at room temperature for 15 minutes. The plates were then incubated at 37°C for 16-18 hours and observed for the zones of inhibition. The growth inhibition zone was measured by a ruler, and results were interpreted as whether the organism was sensitive, intermediate or resistant to the antimicrobial agents by comparison with standard guidelines based on the Clinical and Laboratory Standards Institute 2023 (16).

### Data quality assurance

Different methods were used for assuring data quality. A standard and structured questionnaire prepared in English was used. The questionnaire was then translated to the local language (Afan Oromo) for data collection and then re-translated back into English for analysis. One day of training was given to the data collectors and supervisors on the data collection tool and procedures. To ensure validity, 5% of the questionnaire was pretested. The findings from the pretesting were utilized for modifying and adjusting of the instrument and interviewing technique. The data collectors were supervised closely by the supervisors and the principal investigator. The completeness of each questionnaire was checked daily by the principal investigator and the supervisors. To ensure consistency, coding, double entry and cleaning were performed. The entire data collection process was guided by the principal investigator.

The specimen containers were properly labeled with patient identifier, codes, collector initial and collection date and time. Then, the collected specimens were immediately transported to the clinical microbiology laboratory for processing according to existing SOPs. All patient information was checked for clarity and completeness. Media and all relevant reagents were carefully inspected and checked for expiration dates prior to use. The sterility of the prepared culture media was checked by incubating 5% of the batch at 35–37°C overnight and evaluating it for possible contamination.

Trained laboratory experts performed the tests. All testing procedures were performed depending on the existing SOPs of the clinical microbiology laboratory. *S. aureus* was used for checking the functionality of blood agar and chocolate agar, and *E. coli* (ATCC 25922) and *P. aeruginosa* (ATCC 27853) were used for MacConkey and biochemical tests.

### Antimicrobial susceptibility testing

Antimicrobial susceptibility tests were performed using the Kirby–Bauer disk diffusion method on Muller Hinton agar (MHA) (HIMEDIA, India). Antibiotic discs were selected based on the prescription pattern in the study area and recommendations from the Clinical Laboratory Standards Institute (CLSI). The grades of the susceptibility profile were read as sensitive (S), intermediate (I), or resistant (R) by comparison of the zone of inhibition with clinical and laboratory standards institute guidelines 33^rd^ edition (16). The following antibiotic disks were used for susceptibility testing: Ceftriaxone (30 μg), Cefoxitin (30 μg), Ceftazidime (30 μg), Penicillin G (10 μg), Ampicillin (10 μg), Amoxicillin-clavulanate (20/10μg), Piperacillin-tazobactam (100/10 μg), Meropenem (10 μg), Gentamicin (10 μg), Tobramycin (10 μg), Ciprofloxacin (5 μg), Azithromycin (15 μg), Clindamycin (30 μg), Vancomycin (30 μg), Tetracycline (30 μg), Chloramphenicol (30 μg) and Trimethoprim sulfamethoxazole (1.25/23.75).

### Statistical analysis

The collected data were coded, entered into Epi-data version 4.6.0.6 software and then cleaned. From this software, they were exported to SPSS version 25 for analysis. Descriptive statistics were calculated to describe relevant variables. The data were presented in words, figures, and tables. Binary logistic regression analysis was used to select candidate variables for multivariable logistic regression analysis. Variables with P-values < 0.25 were candidates for multivariate analysis. The adjusted odds ratio (AOR) was used to determine the strength of the association. A P-value <0.05 was considered to indicate statistical significance.

### Ethics approval and consent to participate

This study was approved by the Salale University Institutional Review Board under the number S/U-IRB/25/23. An official permission letter was obtained from Nekemte Public Health Research and Referral Laboratory Center. Participants were informed of the purpose of the study, risks associated with the study, confidentiality of personal data, and their right to take part in the study. After that, we obtained a written informed consent from adult study participants, whereas an assent form was obtained from study participants less than 18 years of age. In addition, a consent was also obtained from their parents or legal guardians to participate in this study. Finally, specimens were collected from all study participants and analyzed accordingly. Laboratory results of study participants were communicated with their respective physicians for better management

## Results

### Characteristics of the study participants

A total of 242 middle ear discharge samples were collected from the study participants and analyzed. Males and females accounted for 129 (53.3%) and 113 (46.7%) of the participants, respectively. The age of the participants ranged from 1 to 65 years, with mean and median ages of 17.1 and 14.0 years, respectively. Ninety-one (37.6%) of them were aged less than five years, while 73 (30.2%) were aged 25 years and older. One hundred sixty-five (68.2%) and 77 (31.8%) of the participants were from urban and rural areas, respectively (Table 1).

**Table 1:** Age, sex and residence distribution of patients with otitis media referred to Nekemte Public Health Research and Referral Laboratory Center, Nekemte, June-September 2023.

| Variable |  | Frequency | Percentage |
| --- | --- | --- | --- |
| Age | <5 | 91 | 37.6 |
|  | 5 – 14 | 34 | 14.0 |
|  | 15 – 24 | 44 | 18.2 |
|  | ≥25 | 73 | 30.2 |
| Sex | Male | 129 | 53.3 |
|  | Female | 113 | 46.7 |
| Residence | Urban | 165 | 68.2 |
|  | Rural | 77 | 31.8 |

### Prevalence of bacterial pathogens

Two hundred twelve middle ear discharge samples were positive for culture resulting in an overall 87.6% prevalence of bacterial isolates. A total of 228 bacterial pathogens were recovered from positive cultures constituting (107, 46.9%) Gram-positive and (121, 53.1%) Gram-negative bacteria. From the total bacterial isolates, *S. aureus* (92, 40.4%) and *P. aeruginosa* (33, 14.5%) were the predominant bacterial species followed by *E. coli* (24, 10.5%) and *K. pneumoniae* (16, 7.0%) (Table 2).

**Table 2:**
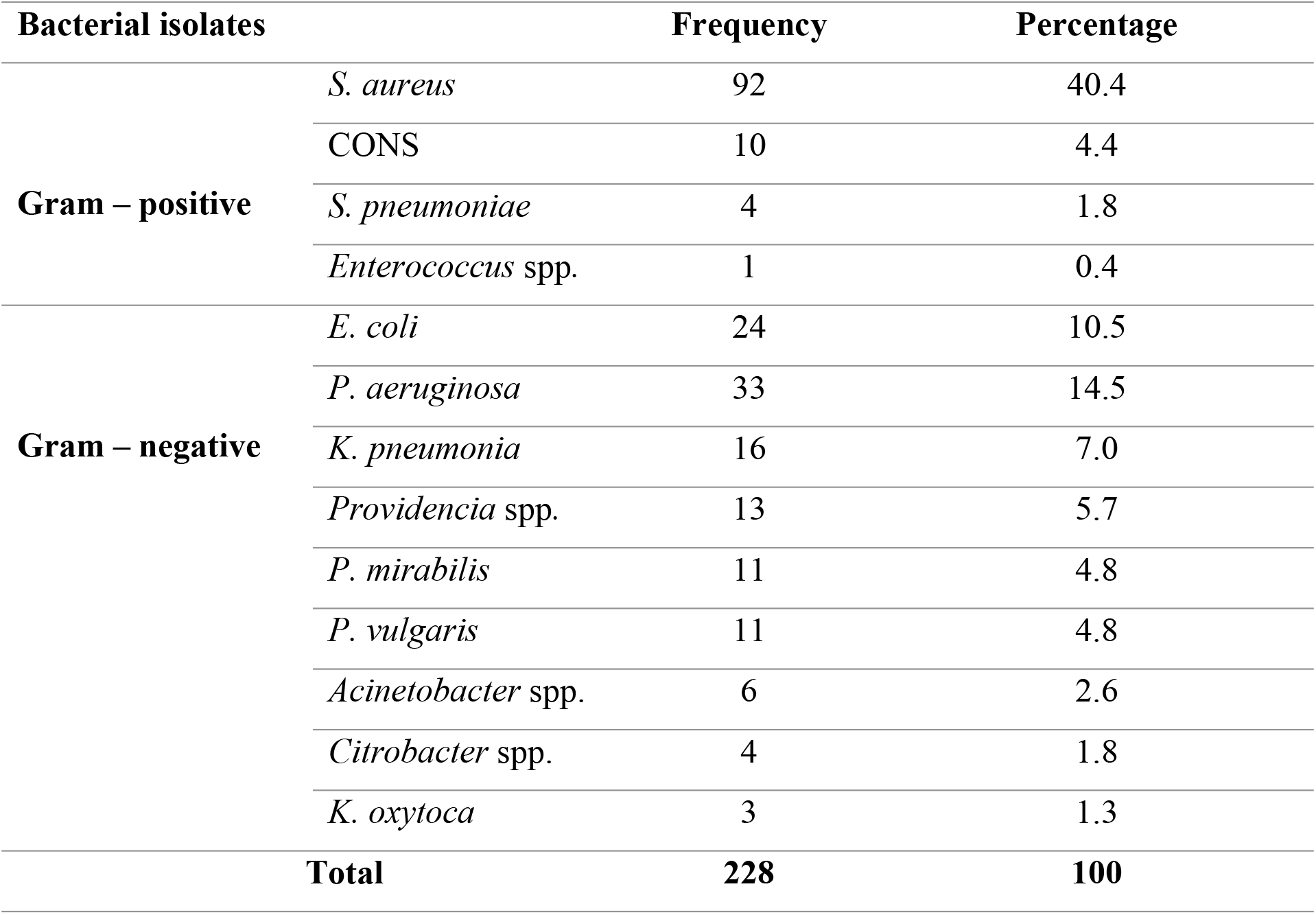
Bacterial etiologic agents among patients with otitis media referred to Nekemte Public Health Research and Referral Laboratory Center, Nekemte, Ethiopia, June-September 2023.

### Antimicrobial susceptibility testing

Antimicrobial susceptibility testing was performed for both gram-positive (n = 107) and gram-negative (n = 121) bacterial pathogens isolated from the study participants. The predominant gram-positive isolate, *S. aureus* showed the highest level of resistance to penicillin (86, 93.5%) followed by cefoxitin (81, 88.0%) but gentamicin (72, 78.3%) and clindamycin (74, 80.4%), the two antibiotics to which *S. aureus* is susceptible (Table 3).

**Table 3:** Antimicrobial resistance patterns of gram-positive bacterial isolates from patients with otitis media referred to Nekemte Public Health Research and Referral Laboratory Center, Nekemte, Ethiopia, June-September 2023.

| Antibiotics tested | Number of resistant bacteria [n (%)] |  |  |  |  |
| --- | --- | --- | --- | --- | --- |
|  | *S. aureus* | CONS | *S. pneumoniae* | *Enterococcus* spp. | Total |
|  | 92 | 10 | 4 | 1 | 107 |
| Ampicillin | NT | NT | NT | 0 (0.0) | 0 (0.0) |
| Azithromycin | 41 (44.6) | 7 (70.0) | 0 (0.0) | NT | 48 (44.9) |
| Cefoxitin | 81 (88.0) | 8 (80.0) | NT | NT | 89 (83.2) |
| Chloramphenicol | 29 (31.5) | 5 (50.0) | NT | 0 (0.0) | 34 (31.8) |
| Ciprofloxacin | 41 (44.6) | 3 (30.0) | NT | NT | 44 (41.1) |
| Clindamycin | 17 (18.5) | 3 (30.0) | 0 (0.0) | NT | 20 (18.7) |
| Cotrimoxazole | 47 (51.1) | 8 (80.0) | 0 (0.0) | NT | 55 (51.4) |
| Gentamicin | 17 (18.5) | 0 (0.0) | NT | NT | 17 (15.9) |
| Penicillin | 86 (93.5) | 8 (80.0) | NT | 0 (0.0) | 94 (87.9) |
| Tetracycline | 71 (77.2) | 9 (90.0) | 2 (50.0) | NT | 82 (76.7) |

CONS – Coagulase negative staphylococci, NT – Not tested

Twenty-three (95.8%), 32 (97%), 16 (100%), and 12 (92.3%) *E. coli, P. aeruginosa*, *K. pneumoniae,* and *Providencia species,* respectively, demonstrated susceptibility to piperacillin-tazobactam. Twenty (60.6%) *P. aeruginosa* strains exhibited resistance to meropenem. The majority of 22 (91.7%) of *E. coli* isolates were resistant to ampicillin (Table 4).

**Table 4:**
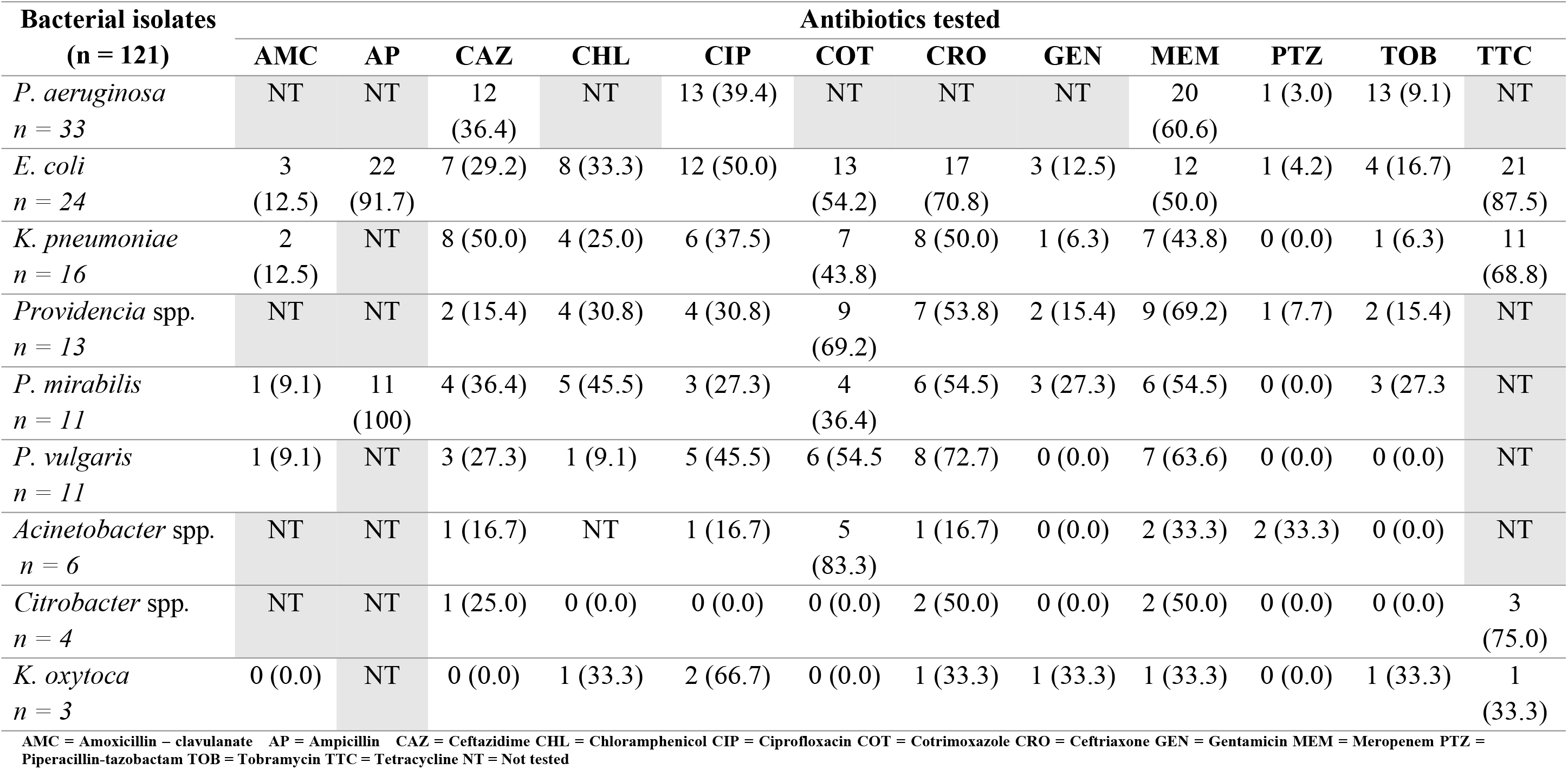
Antimicrobial resistance patterns of gram-negative bacterial isolates from patients with otitis media referred to Nekemte Public Health Research and Referral Laboratory Center, Nekemte, Ethiopia, June-September 2023.

| Bacterial isolates<br>(n = 121) | Antibiotics tested |  |  |  |  |  |  |  |  |  |  |  |
| --- | --- | --- | --- | --- | --- | --- | --- | --- | --- | --- | --- | --- |
|  | AMC | AP | CAZ | CHL | CIP | COT | CRO | GEN | MEM | PTZ | TOB | TTC |
| *P. aeruginosa*<br>n = 33 | NT | NT | 12<br>(36.4) | NT | 13 (39.4) | NT | NT | NT | 20<br>(60.6) | 1 (3.0) | 13 (9.1) | NT |
| *E. coli*<br>n = 24 | 3<br>(12.5) | 22<br>(91.7) | 7 (29.2) | 8 (33.3) | 12 (50.0) | 13<br>(54.2) | 17<br>(70.8) | 3 (12.5) | 12<br>(50.0) | 1 (4.2) | 4 (16.7) | 21<br>(87.5) |
| *K. pneumoniae*<br>n = 16 | 2<br>(12.5) | NT | 8 (50.0) | 4 (25.0) | 6 (37.5) | 7<br>(43.8) | 8 (50.0) | 1 (6.3) | 7 (43.8) | 0 (0.0) | 1 (6.3) | 11<br>(68.8) |
| *Providencia* spp.<br>n = 13 | NT | NT | 2 (15.4) | 4 (30.8) | 4 (30.8) | 9<br>(69.2) | 7 (53.8) | 2 (15.4) | 9 (69.2) | 1 (7.7) | 2 (15.4) | NT |
| *P. mirabilis*<br>n = 11 | 1 (9.1) | 11<br>(100) | 4 (36.4) | 5 (45.5) | 3 (27.3) | 4<br>(36.4) | 6 (54.5) | 3 (27.3) | 6 (54.5) | 0 (0.0) | 3 (27.3) | NT |
| *P. vulgaris*<br>n = 11 | 1 (9.1) | NT | 3 (27.3) | 1 (9.1) | 5 (45.5) | 6 (54.5) | 8 (72.7) | 0 (0.0) | 7 (63.6) | 0 (0.0) | 0 (0.0) | NT |
| *Acinetobacter* spp.<br>n = 6 | NT | NT | 1 (16.7) | NT | 1 (16.7) | 5<br>(83.3) | 1 (16.7) | 0 (0.0) | 2 (33.3) | 2 (33.3) | 0 (0.0) | NT |
| *Citrobacter* spp.<br>n = 4 | NT | NT | 1 (25.0) | 0 (0.0) | 0 (0.0) | 0 (0.0) | 2 (50.0) | 0 (0.0) | 2 (50.0) | 0 (0.0) | 0 (0.0) | 3<br>(75.0) |
| *K. oxytoca*<br>n = 3 | 0 (0.0) | NT | 0 (0.0) | 1 (33.3) | 2 (66.7) | 0 (0.0) | 1 (33.3) | 1 (33.3) | 1 (33.3) | 0 (0.0) | 1 (33.3) | 1<br>(33.3) |

AMC = Amoxicillin – clavulanate AP = Ampicillin CAZ = Ceftazidime CHL = Chloramphenicol CIP = Ciprofloxacin COT = Cotrimoxazole CRO = Ceftriaxone GEN = Gentamicin MEM = Meropenem PTZ = Piperacillin-tazobactam TOB = Tobramycin TTC = Tetracycline NT = Not tested

### Multidrug Resistance

The overall prevalence of MDR bacteria in this study was 151 (66.2%). Among the gram-positive and gram-negative isolates, 91 (39.9%) and 60 (26.3%) were multidrug resistant, respectively. The predominant bacterial pathogens isolates were *S. aureus* (82, 54.3%), *E. coli* (20, 13.2%) and *K. pneumoniae* (11, 7.3%) (Table 5).

**Table 5:** Multidrug resistance patterns of gram-positive and gram-negative bacterial isolates from patients with otitis media referred to Nekemte Public Health Research and Referral Laboratory Center, Nekemte, Ethiopia, June-September 2023.

| Bacteria isolates | Antimicrobial susceptibility test results, No (%) |  |  |  | Total<br>MDR |
| --- | --- | --- | --- | --- | --- |
|  | R0 | R1 | R2 | ≥R3 |  |
| Gram-positive isolates |  |  |  |  |  |
| *S. aureus* (n = 92) | 0 (0.0) | 5 (5.4) | 5 (5.4) | 82 (89.1) | 82 (76.6) |
| *CONS* (n = 10) | 0 (0.0) | 1 (10.0) | 0 (0.0) | 9 (90.0) | 9 (8.4) |
| *S. pneumoniae* (n = 4) | 3 (75) | 1 (25.0) | 0 (0.0) | 0 (0.0) | 0 (0.0) |
| *Enterococcus* spp. (n = 1) | 1 (100) | 0 (0.0) | 0 (0.0) | 0 (0.0) | 0 (0.0) |
| **Total (n = 107)** | **4 (3.7)** | **7 (6.5)** | **5 (4.7)** | **91 (85.1)** | **91 (85.1)** |
| Gram-negative isolates |  |  |  |  |  |
| *P. aeruginosa* (n = 33) | 6 (18.2) | 11 (33.3) | 11 (33.3) | 5 (15.2) | 5 (4.1) |
| *E. coli* (n = 24) | 0 (0.0) | 2 (8.3) | 2 (8.3) | 20 (83.3) | 20 (16.5) |
| *K. pneumoniae* (n = 16) | 1 (6.3) | 0 (0.0) | 4 (25.0) | 11 (68.8) | 11 (9.1) |
| *Providencia* spp. (n = 13) | 2 (15.4) | 2 (15.4) | 2 (15.4) | 7 (53.8) | 7 (5.8) |
| *P. mirabilis* (n = 11) | 0 (0.0) | 0 (0.0) | 3 (27.3) | 8 (72.7) | 8 (6.6) |
| *P. vulgaris* (n = 11) | 1 (9.1) | 1 (9.1) | 3 (27.3) | 6 (54.5) | 6 (5.0) |
| *Acinetobacter* spp. (n = 6) | 1 (16.7) | 1 (16.7) | 2 (33.3) | 2 (33.3) | 2 (1.7) |
| *Citrobacter* spp. (n = 4) | 4 (100) | 0 (0.0) | 0 (0.0) | 0 (0.0) | 0 (0.0) |
| *K. oxytoca* (n = 3) | 1 (33.3) | 1 (33.3) | 0 (0.0) | 1 (33.3) | 1 (0.8) |
| **Total (n = 121)** | **16 (13.2)** | **18 (14.9)** | **27 (22.3)** | **60 (49.6)** | **60 (49.6)** |

R0 - ≥R3 refers to the number of pathogenic bacterial isolates resistant to 0 - ≥3 different antibiotics

### Possible risk factors for otitis media

Both bivariate and multivariate logistic regression analyses were performed to assess the possible risk factors for middle ear infection. Statistical significance was obtained for purulent middle ear discharge, duration of middle ear discharge ≥14 days and history of active/passive smoking. Study participants with purulent middle ear discharge were approximately six times more likely to have positive cultures for bacterial pathogens responsible for causing otitis media [AOR = 6.534 (95% CI: 2.112 – 20.208; P-value = 0.001)]. On the other hand, participants who experienced otorrhea of ≥14 days are approximately seven times more likely to have positive cultures than were those who experienced otorrhea of <14 days [7.628 (95% CI = 3.135 – 18.558; P-value = 0.000)]. Those who had an active or passive smoking history were approximately eight times more likely to develop otitis media than to those without a history of active or passive smoking [8.817 (95% CI = 1.072 – 72.534; P-value = 0.043)] (Table 6).

**Table 6:** Multivariate analyses to identify associated factors among study participants with otitis media referred to Nekemte Public Health Research and Referral Laboratory Center, Nekemte, June – September 2023.

| Variables |  | Culture Result, No (%) |  | COR (95% CI) | AOR (95% CI) | P-value |
| --- | --- | --- | --- | --- | --- | --- |
|  |  | Positive | Negative |  |  |  |
| Age group | <5 | 75 (31.0) | 16 (6.6) | 0.272 (0.087 – 0.853) | 0.436 (0.116 – 1.641) | 0.220 |
|  | 5 – 14 | 28 (11.6) | 6 (2.5) | 0.271 (0.071 – 1.032) | 0.524 (0.112 – 2.455) | 0.412 |
|  | 15 – 24 | 40 (16.5) | 4 (1.7) | 0.580 (0.137 – 2.446) | 0.672 (0.132 – 3.408) | 0.631 |
|  | ≥25 | 69 (28.5) | 4 (1.7) | **Reference** | **Reference** |  |
| Residence | Rural | 71 (29.3) | 6 (2.5) | 2.014 (0.788 – 5.151) | 0.923 (0.305 – 2.791) | 0.880 |
|  | Urban | 141 (58.3) | 24 (9.9) | **Reference** | **Reference** |  |
| Ear involved | Right | 114 (47.1) | 20 (8.3) | **Reference** | **Reference** |  |
|  | Left | 98 (40.5) | 10 (4.1) | 1.719 (0.768 – 3.848) | 1.219 (0.470 – 3.162) | 0.683 |
| Appearance of middle ear discharge | Bloody | 17 (7.0) | 9 (3.7) | **Reference** | **Reference** |  |
|  | Purulent | 157 (64.9) | 10 (4.1) | 8.312 (2.967 – 23.288) | **6.534 (2.112 – 20.208)** | **0.001\*** |
|  | Mucoid | 38 (15.7) | 11 (4.5) | 1.829 (0.640 – 5.228) | 1.398 (0.427 – 4.580) | 0.580 |
| Duration of middle ear discharge | <14 days | 34 (14.0) | 18 (7.4) | **Reference** | **Reference** |  |
|  | ≥14 days | 178 (73.6) | 12 (5.0) | 7.853 (3.468 – 17.783) | **7.628 (3.135 - 18.558)** | **0.000\*** |
| History of smoking | No | 164 (67.8) | 29 (12.0) | **Reference** | **Reference** |  |
|  | Yes | 48 (19.8) | 1 (0.4) | 8.488 (1.127 – 63.936) | **8.817 (1.072 – 72.534)** | **0.043\*** |
| Treatment history for otitis media | No | 122 (50.4) | 21 (8.7) | **Reference** | **Reference** |  |
|  | Yes | 90 (37.2) | 9 (3.7) | 1.721 (0.753 – 3.936) | 0.895 (0.354 – 2.736) | 0.976 |

\*Associated factors for otitis media
CI = Confidence interval, COR = Crude odds ratio, AOR = Adjusted odds ratio

## Discussion

Otitis media is a major reason people seek medical attention globally, and its complications play a significant role in the development of preventable hearing loss, particularly in developing nations (17). According to the present study OM was found to be a common health problem at all ages. According to the present study, OM was found a common health problem in all ages. However, a peak prevalence of 31.0% was observed among children under five years of age, which was similar to that reported in other studies in Ethiopia (28.1%) (18), higher than that reported in India (14.9%) (19) but lower than that study performed in Yemen (66.7%) (20). This prevalence of OM in children is mainly attributed to immature immune status, the anatomy of the Eustachian tube (shorter and horizontal), frequent exposure to URTIs and malnutrition.

Gender-wise analysis of this study showed that males were more affected than females were. This finding, with male predominance, is in agreement with a study performed in Wollo, Ethiopia (50.4%) (13) and Pakistan (43.9%) (21) but lower than a study performed in India (56.7%) (22). In contrast, other studies in Iran (57.8%) and Iraq (60.0%) (23,24) showed that females were more affected by otitis media than males were. The differences in male and female predominance may be attributed to the nature of the sampling technique.

This study also provides insight into the prevalence of otitis media with respect to the residential location of study participants. The prevalence of otitis media among study participants from urban areas was 58.3%. This finding agrees with a study performed in Mekele, Ethiopia (52.0%) (25) but is lower than that in a study performed in Gondar, Ethiopia (76.5%) (26). However, the present study disagrees with a study performed in China (27) in which 85.9% of rural areas were positive for OM. These disparities may come from the involvement of study participants from urban areas due to increased health seeking-behavior, proximity to health facilities and culture diagnostic services in the study areas.

In the present study, the overall middle ear discharge culture positivity rate was 87.6% (95% CI = 82.8 – 91.5). This finding is similar to that of study performed in Dessie, Ethiopia, which reported 89.4% (28) but it was higher than that of studies done in Gondar (76.7%) and Bahir Dar (80.4%) (14,29). In contrast, the current culture positivity rate was lower than that studies performed in Ghana (97%) and India (95.7%) (30,31), which may be related to variations in the availability of isolation and identification media. This may be correlated with the fact that availability of relevant media and other supplies maximizes the frequency of culture positivity in middle ear discharge.

A total of 228 bacterial isolates were identified. Analysis of the Gram reactions of the isolates revealed that 53.1% [95% CI (46.6 – 59.5)] and 46.9% [95% CI (40.5 – 53.4%)] were gram-negative and gram-positive bacterial pathogens, respectively. Another study in Ethiopia reported 56.0% gram-negative bacteria as the predominant species isolated from middle ear discharge, which was consistent with the results of the present study (25). Reports from Somalia, Nigeria and Malaysia agree with the predominance of gram-negative bacteria, with higher frequencies of 77.3%, 71.6% and 75.3%, respectively (32–34). The reason for the higher prevalence could be the chronic nature of infection, where gram-negative bacteria from external sources gain access to the auditory canal and eventually become predominant.

The predominant bacterial isolates in this study were *S. aureus* (40.5%) and *P. aeruginosa* (14.5%) which was similar to the findings of studies in Ethiopia (35), Pakistan (36) and China (37) but inconsistent with the findings of other studies in Ethiopia (13), India (38) and Turkey (39). In addition, the isolation rates of *Coagulase negative staphylococci*, *S. pneumoniae* and *Enterococcus species* were 4.4%, 1.8% and 0.4%, respectively. At least one of these bacterial isolates has also been reported in other studies (25,26,29,40–42). This study revealed that gram-negative bacterial isolates included *E. coli* (19.8%), *K. pneumoniae* (13.2%)*, Providencia species* (10.7%), *P. mirabilis* (9.1%), *P. vulgaris* (9.1%), *Acinetobacter species* (5.0%), *Citrobacter species* (3.3%) and *K. oxytoca* (2.5%). Other studies from Ethiopia and other countries have also reported these bacteria at varying frequencies (8,13,18,26,33,43). Variations in climate and geography are possible reasons for the differences in the distribution of bacterial isolates (29).

In the present study, 93.5% of the *S. aureus* isolates resistant to penicillin whereas clindamycin and gentamicin were the two drugs to which 80.4% and 78.3% of isolates were susceptible, respectively. This finding is in agreement with other studies (29,32). On the other hand, *P. aeruginosa* demonstrated susceptibility patterns ranging from 33.3 – 97% with piperacillin-tazobactam being the most effective antibiotic but 60.6% of the isolates were found to be resistant to meropenem. Studies from other parts of Ethiopia used different antibiotics and reported varying susceptibility patterns (25,35). Studies from other parts of the world tested different antibiotics with varying susceptibility patterns (30,36,44) with one of the studies (30) that tested piperacillin-tazobactam, ciprofloxacin and ceftazidime, which were also tested in this study. The use of different antibiotics in different studies is attributed to the occasional emergence of resistant strains from time to time, the availability of proposed antibiotics and local prescribing practices.

The overall MDR rate in this study was 66.2% [95% CI (59.7 – 72.3%)]. This finding is similar to those of studies performed in Ethiopia, which reported MDR rates of 67.0% (15) and 61.5% (45), but lower than those of another study that reported rates of 88.3% (26), however; these rates are higher than those of a study performed in India (46). Variations in the reports of MDR rates might be attributed to differences in operational definitions of MDR strains, bacterial isolates and antibiotic discs tested.

There were statistically significant differences in culture positivity between those who were with purulent middle ear discharge compared to those who presented with other middle ear discharge appearances [p-value = 0.001, AOR = 6.534 (95% CI: 2.112 – 20.208). In contrast, a study done in Jimma reported that there was no significant association between purulent discharge and culture positivity (15), but one study from Iraq revealed a statistically significant association between middle ear discharge culture positivity and purulent discharge (44).

A significant difference was also observed between a middle ear discharge duration of ≥14 days and culture positivity [p-value = 0.000, AOR = 7.628 (95% CI: 3.135– 18.558)], which was also supported by other studies (25,26). Chronic otitis is characterized by middle ear discharge that lasts for at least 14 days and a perforated tympanic membrane. Another risk factor for otitis media was a history of smoking in either an active or passive state. A significant association was observed between patients with history of smoking and the development of otitis media [p-value = 0.043, AOR = 8.817 (95% CI: 1.072 – 72.534)]. This association was also supported by other studies (15,47,48). Smoking decreases the mucociliary activity of the respiratory epithelium, depresses local immune function, and enhances the adhesion of bacteria to the respiratory epithelium (48).

### Strengths and limitations of the study

The strength of the study was that all the laboratory procedures were conducted following standard operating procedures. In addition, further studies can be built upon these findings, as there are no published data regarding the problem in the study area. The temporal relation between the exposure and outcome variables could not be established because the study design was cross-section in nature. The small sample size and convenience sampling nature of this study prevented it from being representative of patients with otitis media in Ethiopia. On the other hand, anaerobic culture methods for fastidious bacteria and molecular techniques were not employed.

### Conclusion

The present study indicated that bacterial middle ear infection has become an increasing health problem coupled with levels of multidrug resistance. *S. aureus* and *P. aeruginosa* were the leading causes for middle ear infection. There is an increase in the number of antibiotic-resistant bacteria recovered from patients with otitis media in the study area, and these bacteria are becoming a major public health problem in the management of patients with middle ear infection.

## Data Availability

All data produced in the present work are contained in the manuscript

## Abbreviations

AOM: acute otitis media
AOR: adjusted odds ratio
ATCC: American Type Culture Collection
CI: confidence interval
CLSI: Clinical Laboratory Standards Institute
CSOM: chronic suppurative otitis media
MDR: multidrug resistance
MHA: Muller Hinton Agar
OM: otitis media
SOP: standard operating procedure
SPSS: statistical software for social sciences
URTI: upper respiratory tract infection

## Acknowledgements

The authors are grateful to Salale University and the Oromia Health Bureau for providing opportunity to conduct this research. We also acknowledge Nekemte Public Health Research and Referral Laboratory Center for providing the laboratory setup, chemicals and reagents. Our deepest gratitude also goes to the study participants, staff members of the Clinical Microbiology Department of Nekemte Public Health Research and Referral Laboratory Center, referring health facilities and data collectors and supervisors. We would also sincerely thank the study participants for their participation in the study.

## Authors’ contributions

**Conceptualization:** Endalu Tesfaye Guteta, Fedasan Alemu Abdi, Tadesse Bekele Tafesse

**Data curation**: Endalu Tesfaye Guteta, Fedasan Alemu Abdi, Seifu Gizaw Feyisa, Tadesse Bekele Tafesse

**Formal analysis**: Endalu Tesfaye Guteta, Fedasan Alemu Abdi, Tadese Bekele Tafesse

**Funding acquisition**: Endalu Tesfaye Guteta, Fedasan Alemu Abdi, Belay Merkeb Zewudie

**Investigation**: Endalu Tesfaye Guteta, Betrearon Sileshi Kinfu, Hunduma Feyisa Geleta

**Methodology**: Endalu Tesfaye Guteta, Seifu Gizaw Feyisa, Fedasan Alemu Abdi, Belay Merkeb Zewudie, Betrearon Sileshi Kinfu, Hunduma Feyisa Geleta, Tadesse Bekele Tafesse

**Project administration**: Endalu Tesfaye Guteta, Seifu Gizaw Feyisa, Fedasan Alemu Abdi, Belay Merkeb Zewudie, Betrearon Sileshi Kinfu, Hunduma Feyisa Geleta, Tadesse Bekele Tafesse

**Resources**: Endalu Tesfaye Guteta

**Software**: Endalu Tesfaye Guteta

**Supervision**: Endalu Tesfaye Guteta, Fedasan Alemu Abdi, Betrearon Sileshi Kinfu, Hunduma Feyisa Geleta

**Validation:** Endalu Tesfaye Guteta, Fedasan Alemu Abdi, Tadesse Bekele Tafesse

**Visualization**: Endalu Tesfaye Guteta, Fedasan Alemu Abdi, Tadese Bekele, Seifu Gizaw Feyisa

**Writing – original draft**: Endalu Tesfaye Guteta

**Writing – review and editing**: Endalu Tesfaye Guteta, Seifu Gizaw Feyisa, Fedasan Alemu Abdi, Belay Merkeb Zewudie, Tadesse Bekele Tafesse

## Data Availability Statement

All relevant data are within this paper

